# Navigating Gestational Health in the Reddit Pregnancy Community: A Mixed-Methods Study of Online Peer Discourse and Embodied Experience

**DOI:** 10.64898/2026.11.24.26368774

**Authors:** Luba Marderfeld, Jordyn M. Cox, Taniya Nagpal, Lavy Khoushinsky, Kristi Adamo

## Abstract

**Background:** While health behaviours and clinical markers such as gestational weight gain GWG), physical activity, and nutrition are critical for optimizing maternal and fetal outcomes, pregnant individuals often face a fragmented informational landscape characterized by a heavy burden of clinical restrictions. Consequently, many turn to online, pseudo-anonymous communities like Reddit to crowdsource support, share experiences, and navigate complex health decisions outside the formal clinical encounter. This study aimed to examine how pregnant individuals utilize the Reddit community r/Pregnant to seek health-related information, navigate clinical recommendations surrounding GWG and health-related behaviours, and share lived experiences.

**Methods:** An explanatory sequential mixed-methods study design was employed, combining computational topic modeling and qualitative analysis. In aim 1, all public posts published on the r/Pregnant subreddit between 2009 and 2024 were analyzed using BERTopic, a transformer-based modeling framework, to map the landscape of discourse and identify health-related topics. In aim 2, nine health-related topics were purposively selected, and 700 high-engagement posts (ranked by comment volume) across topics were analyzed using reflexive thematic analysis, following Braun and Clarke’s six-phase framework.

**Results:** BERTopic identified 90 distinct pregnancy-related topics across 15 years of discourse. Nine were health-related and selected for analysis, with gestational weight gain having the highest volume (Rank 14; 2,241 posts). Reflexive thematic analysis generated five overarching themes: (1) Reddit as a Site of Collective Sense-Making; (2) Decision-Making Under Uncertainty; (3) The Emotional Burden of Maternal Responsibility; (4) Embodied Knowledge in Practice; and (5) Social Pressure and Autonomy.

**Conclusions:** A substantial gap exists between standardized prenatal clinical guidelines and the lived, physical reality of pregnancy. Pregnant individuals actively use digital peer support to adapt clinical advice, cope with weight-related stigma, and exercise autonomy. These findings highlight the importance of strengthening prenatal healthcare communication through more empathetic, non-judgmental, and individualized approaches that better reflect pregnant individuals’ concerns and lived experiences.

## Background

Health behaviours during pregnancy, like physical activity (PA), dietary intake, sleep, and prenatal care management, represent key modifiable factors that influence maternal and infant outcomes, as well as physiological parameters such as gestational weight gain (GWG) (Hill et al., 2021; Lu et al., 2021; McElfish et al., 2026; “ACOG,” 2020; Teede et al., 2022). Although GWG itself is an outcome of complex physiological and behavioural processes rather than a health behaviour, it serves as a clinical marker monitored throughout antenatal care (American College of Obstetricians and Gynecologists Committee on Clinical Consensus-Obstetrics, 2025).

The clinical importance of optimizing GWG through health behaviours is well established. Insufficient GWG elevates the risk of small-for-gestational-age infants and preterm birth (Goldstein et al., 2017). Whereas, excessive GWG significantly increases the odds of fetal macrosomia, caesarean delivery, gestational diabetes, and hypertensive disorders of pregnancy, includingpreeclampsia. (Goldstein et al., 2017; Khan et al., 2023). Conversely, regular moderate-intensity exercise lowers the risk of excessive GWG, preeclampsia, gestational diabetes, and caesarean birth in uncomplicated pregnancies (Barone Gibbs et al., 2026; Mottola et al., 2018; “ACOG,” 2020).

To promote optimal gestational outcomes, clinical guidelines recommend accumulating at least 150 minutes of moderate-intensity PA weekly (Evenson et al., 2019; Mottola et al., 2018; “ACOG,” 2020), which facilitates achieving GWG targets recommended by the Institute of Medicine (IOM) (*Weight Gain During Pregnancy*, 2009). Despite these evidence-based recommendations, adherence remains poor: only about one-third of pregnant women achieve GWG within recommended ranges, and as few as one in four meet PA guidelines (Barone Gibbs et al., 2026; Creanga et al., 2022). Unhealthy dietary behaviours and higher energy intake are associated with greater GWG and have also been linked to gaining weight outside the recommendations, although evidence for nutrition knowledge itself is inconsistent (Kapadia et al., 2015; Souza et al., 2020; You et al., 2023). Additional data illustrate that eating behaviours, such as food cravings and, in some studies, restrained eating, are associated with higher GWG (Bijlholt et al., 2020; Orloff et al., 2016).

Beyond PA and diet, pregnant women also face challenges related to sleep: disturbed sleep is more common than in non-pregnant women, and sleep duration declines from the second trimester onward, with the proportion of short sleepers rising sharply in late pregnancy (Aukia et al., 2024; Plancoulaine et al., 2017). Shorter sleep during pregnancy is associated with higher blood glucose and an increased risk of gestational diabetes (Facco et al., 2017).

Clinical guidelines also recommend a decrease in maternal caffeine intake, generally restricting daily consumption to a maximum threshold of 200 to 300 mg (Anderson et al., 2009; Jin & Qiao, 2021). Elevated maternal caffeine consumption is associated with adverse outcomes such as low birth weight, preterm birth, and pregnancy loss; notably, the odds of low birth weight increase by approximately 3% for each additional 100 mg consumed daily (Jin & Qiao, 2021). Furthermore, pregnant individuals are advised to avoid high-risk sources of *Listeria* contamination (such as unpasteurized dairy products, soft cheeses, and deli meats), because an estimated 95% of maternal listeriosis cases result in severe adverse outcomes, including fetal loss, preterm delivery, and neonatal infection (“Committee Opinion No. 614,” 2014; Disson et al., 2025).

Prenatal guidance extends to nutritional supplementation as well, for example, 400 µg of folic acid daily is recommended for all women who are planning a pregnancy or could become pregnant, to reduce the risk of neural tube defects (NTDs) (US Preventive Services Task Force et al., 2023). Iron, iodine, vitamin D and omega-3 (DHA) are also commonly recommended, but the evidence that they improve outcomes is weaker, and guidelines disagree on them (“Anemia in Pregnancy,” 2021; Bath, 2024; Demay et al., 2024). In Canada, women are advised to take a daily multivitamin with 400 µg folic acid from before conception through breastfeeding (Lamers et al., 2018). Adherence is high once women are pregnant but poor before conception, and the doses women take are often wrong (Hopperton et al., 2025).

Navigating this extensive matrix of clinical recommendations and lifestyle restrictions places a significant burden on pregnant individuals (Lupton, 2012). Public understanding of health behaviours remains fragmented and is increasingly shaped by sources outside the clinical encounter. Pregnant individuals routinely source information on exercise, nutrition, and GWG from digital platforms such as Facebook, Instagram, and TikTok; an estimated 97.5% use the internet for pregnancy-related information, and 89% engage on social media (Baker & Yang, 2018). Because unregulated platforms can propagate misinformation and unrealistic portrayals of pregnancy, including anecdotal advice from ‘fitness influencers’, the information women encounter may diverge from evidence-based guidance (Djouma Nembot et al., 2026; Hayman et al., 2023). Internet use for pregnancy information is longstanding: a systematic review of studies published through 2014 found that most pregnant women sought such information online, at least monthly, with fetal development and nutrition among the topics searched most frequently (Sayakhot & Carolan-Olah, 2016). Online spaces are appealing because they offer continuous access, peer comparison, and a setting to voice concerns outside formal clinical structures (Asadollahi et al., 2026; Hussain et al., 2023; Snyder et al., 2020).

Among these online spaces, discussion forums are particularly relevant because they combine information seeking with experiential and emotional support. However, peer-generated content may expose users to incomplete, contradictory, or clinically unsafe advice. In a forum analysis by Ellis & Roberts (Ellis & Roberts, 2020), only 601 of 1,098 responses sharing advice were judged accurate, and poor-quality contributions were rarely corrected. Furthermore, many pregnant women do not discuss internet-derived information with healthcare providers (Sayakhot & Carolan-Olah, 2016), even though internet content directly influences health decisions regarding delivery, medication, and exercise (Taştekin Ouyaba & İnfal Kesim, 2021).

Reddit is a prominent example of such a forum platform: it is the third most-visited website in Canada, averaging 301 million monthly visits (*Canada Leading Websites by Monthly Traffic 2024*, n.d.). The subreddit r/pregnant hosts 1.7 million members who share experiences, ask for support, and discuss gestational health. Reddit’s pseudo-anonymous nature provides an ‘incognito’ environment where individuals share sensitive health details they might hesitate to disclose in person. Such messages are in the form of free text that can be analyzed using natural language processing (NLP), which has been largely driven by the development of pre-trained language models such as Bidirectional Encoder Representations from Transformers (BERT) (Vaswani et al., 2017). One methodology that builds on these advances is BERTopic (Grootendorst, 2022), which uses language embeddings and clustering techniques to extract topics from large volumes of text, a technique that has been successfully applied across a wide range of use cases (Kokkinakis & Hammarlin, 2024).

However, previous studies examining digital pregnancy discourse have been limited by small, geographically restricted samples and heterogeneous methods (Dalhaug & Haakstad, 2019). No study to date has analyzed the complete post corpus of a dedicated online pregnancy community such as r/pregnant. Applying BERTopic to this full corpus therefore offers an opportunity to identify at scale which health-related topics pregnancy individuals discuss and how they make sense of them, in a setting where peer exchange sits alongside, and may diverse from, evidence-based guidance.

### Study Aims

This study utilized an explanatory equential mixed-methods design to explore how pregnant individuals utilize an online community to navigate, discuss, and make sense of gestational weight gain (GWG), health-related behaviours, and gestational outcomes throughout pregnancy. To achieve this, the study pursued two specific, sequential aims:

**Aim 1:** To computationally map the macro-level landscape of prenatal discourse across a 15-year dataset to identify the topics of concern.

**Aim 2:** To utilize the computationally identified topics from Aim 1 to purposively sample highly engaged discussions, qualitatively exploring the nuanced socio-cultural narratives and collective sensemaking processes surrounding gestational weight gain and related prenatal health behaviours.

## Methods

### Study Design Overview

An explanatory sequential mixed-methods design was utilized. Aim 1 involved a computational topic modeling pipeline to map the macro-level discourse of the entire dataset and identify health-related topics. Aim 2 utilized these computational outputs to purposively sample highly engaged discussions for an in-depth, qualitative reflexive thematic analysis.

### Topic Modeling & Computational Pipeline

All public posts published on the r/pregnant subreddit between 2009 and 2024 were collected and analyzed using BERTopic (Grootendorst, 2022), a modern topic modeling framework integrating transformer-based language models with class-based Term Frequency–Inverse Document Frequency (c-TF-IDF) (Devlin et al., 2019; Kokkinakis & Hammarlin, 2024; Ramos, 2003).

Data preprocessing removed duplicates, URLs, user mentions, and formatting artifacts while preserving textual context. Each post was converted into a dense vector representation using a pre-trained sentence-transformer model (all-MiniLM-L6-v2). Dimensionality reduction was performed using Uniform Manifold Approximation and Projection (UMAP)(McInnes et al., 2018), followed by clustering via Hierarchical Density-Based Spatial Clustering of Applications with Noise (HDBSCAN) to group semantically similar posts and segregate noise (McInnes et al., 2017). Topic term weighting was computed using c-TF-IDF, and representations were reviewed and labeled based on top-weighted terms and representative posts.

### Qualitative Study Design & Sampling

Following BERTopic topic generation, the interdisciplinary research team reviewed the 90 resulting topics to identify those central to maternal health behaviours. To assess clinical relevance and alignment with the study’s research aims, the team evaluated each topic based on its direct connection to modifiable prenatal health factors (e.g., dietary intake, physical activity, and sleep) and psychosocial elements known to influence energy balance, GWG trajectories, and gestational outcomes as outlined in standard prenatal clinical guidelines.

Through this collaborative evaluation process, nine topics (out of 90 total topics, Suppl. Table 1) were purposively selected for qualitative analysis: Weight gain, Prenatal supplements, Cravings, Prenatal exercise, Hunger, Caffeine, Prenatal sleep, Anxiety, and Food safety. For each chosen topic, the highest-engagement posts (ranked by comment volume) were extracted. This mixed-methods approach allowed us to computationally identify the community’s health-related concerns and extract a targeted sample of highly engaged posts, while the subsequent qualitative analysis facilitated an in-depth exploration of the nuanced narratives driving those discussions.

### Reflexive Thematic Analysis

Qualitative data were analyzed using reflexive thematic analysis within an interpretive paradigm (Braun & Clarke, 2006, 2022, 2024). The analysis adopted an inductive, data-driven approach, utilizing semantic coding within an experiential orientation. Analysis followed Braun and Clarke’s six-phase framework: (1) data familiarization, (2) initial coding, (3) generating themes, (4) reviewing themes, (5) defining and naming themes, and (6) producing the report. Data were coded in NVivo 15 by a dietitian (LM), who developed initial themes that were subsequently refined through collaborative reflexive engagement with a women’s health researcher, involving analytic meetings, challenging individual interpretations, and reworking theme boundaries (JMC).

### Reflexivity & Team Positionality

Continuous reflexivity was maintained throughout the research process through the use of a reflexive journal and team analytic discussions. The interdisciplinary research team comprised a doctoral student and registered dietitian (LM), a doctoral student in kinesiology and women’s health (JMC), and health scientists (KA, TN) with expertise across preconception, pregnancy, and postnatal periods. Through ongoing reflexive discussions, the team critically examined how their distinct professional lenses shaped data interpretation; for example, the primary coder’s dietetic background sensitized the analysis to nuances in nutrition and supplementation, while the wider team’s expertise in women’s health and weight stigma attuned the interpretation to themes of maternal surveillance and bodily autonomy.

### Ethics and Data Management

Data collection adhered strictly to public social media research ethics guidelines. As this study relied exclusively on publicly available information with no reasonable expectation of privacy, it was exempt from Research Ethics Board (REB) review in accordance with Article 2.2 of the Tri-Council Policy Statement (TCPS2). All direct identifiers, including Reddit usernames, were permanently excluded to protect user privacy.

## Results

### Computational Topic Prevalence (BERTopic)

BERTopic generated 90 distinct thematic clusters across 15 years of r/pregnant discourse. (Suppl. Table 1) Feeling fetal movement emerged as the single most discussed topic overall (Rank 1; 5,454 posts). Gestational weight gain was the most prevalent health-related topic (Rank 14; 2,241 posts). Together, the nine health-related topics of interest accounted for over 10,000 posts (Table 1).

**Table 1:** Nine Selected Health-Related BERTopic Clusters Analyzed Qualitatively.

| **Topic Rank** | **Topic Name** | **Total Posts (N)** | **Posts Analyzed (n)** | **Representative c-TF-IDF Keywords** |
|---|---|---|---|---|
| 14 | Gestational weight gain | 2,241 | 150 | weight, gain, gained, lbs, weight gain, pounds, gaining |
| 18 | Prenatal supplements | 1,894 | 50 | iron, prenatal, vitamins, vitamin, taking, prenatals, folic acid |
| 34 | Cravings | 1,318 | 50 | cravings, craving, food, eat, what, to eat |
| 35 | Prenatal exercise | 1,315 | 100 | exercise, yoga, workout, pregnancy, do, pregnant |
| 63 | Hunger | 869 | 100 | hungry, eat, appetite, eating, hunger, food |
| 64 | Caffeine | 868 | 50 | coffee, caffeine, tea, drink, cup, drinking |
| 84 | Prenatal sleep | 624 | 100 | sleeping, sleep, side, back, sleeping on back |
| 85 | Prenatal anxiety | 607 | 50 | anxiety, panic, fear, worrying, anxious |
| 89 | Food safety | 591 | 50 | listeria, eat, deli meat, sandwich, cheese, safe |

Note. N = total number of posts identified within the cluster; n = subset of posts purposively sampled for reflexive thematic analysis; c-TF-IDF = class-based term frequency-inverse document frequency (the algorithm used to extract the most representative words for each topic).

### Qualitative Synthesis: Overarching Themes and Sub-Themes

A reflexive thematic analysis of 700 high-engagement posts was conducted to capture complex subjective experiences that BERTopic’s semantic clustering inherently misses. The 700 posts across 9 topics, selected based on the highest number of comments, were published between 2017 and 2024.

Because these concepts recurred consistently across all nine health topics generated by BERTopic, the results were synthesized into a single framework, yielding five overarching themes and 16 sub-themes characterizing digital peer discourse and the embodied pregnancy experience (Table 2).

**Table 2:** Overarching Themes and Sub-Themes Generated from Reflexive Thematic Analysis.

|  |  |
|---|---|
| **1. Reddit as a Site of Collective Sense-Making** | 1.1 Seeking Reassurance, Validation, and Normalization |
|  | 1.2 Exchanging Lived Experience as Peer Knowledge |
|  | 1.3 Reciprocal Encouragement and Gratitude |
| **2. Decision making under uncertainty** | 2.1 Seeking Information Beyond Healthcare |
|  | 2.2 Whose Expertise Counts? |
|  | 2.3 Conflicting and Contradictory Guidance |
|  | 2.4 Balancing pregnancy guidance with personal needs and perceived risk |
| **3. The Emotional Burden of Maternal Responsibility** | 3.1 Fear and Anxiety Around Pregnancy |
|  | 3.2 Guilt, Self-Blame, and Maternal Responsibility |
|  | 3.3 Body Image and Weight-Related Identity |
| **4. Embodied Knowledge in Practice** | 4.1 The Body's Unpredictability: Hunger, Appetite, and Physical Extremes |
|  | 4.2 The Shifting Palate |
|  | 4.3 Adapting Recommendations to Lived Reality |
| **5. Social Pressure and Autonomy** | 5.1 External and Intimate Surveillance |
|  | 5.2 Provider Judgment and Pushback |
|  | 5.3 Resistance, Autonomy, and Reclaiming Agency |

### Theme 1: Reddit as a Site of Collective Sense-Making

Pregnant individuals utilized Reddit to compare experiences, normalize physical challenges, and find non-judgmental support.

#### 1.1 Seeking Reassurance, Validation, and Normalization

Sub-theme 1.1 highlights how pregnant individuals use Reddit for emotional regulation, leveraging peer experiences to validate choices, normalize anxieties, and mitigate fear. The discourse features a strong need for reassurance, a desire for non-judgmental spaces, and the sharing of negative emotions related to missed supplements, unexpected physical symptoms and bodily changes, such as weight gain.*“Someone else PLEASE tell me you’re also basically a bottomless pit so I can feel less alone”* (Hunger, 2019) and *“Someone please tell me they got unexpectedly pregnant and didn’t take vitamins until they found out and their baby was/is okay”* (Supplements, 2023).

#### 1.2 Exchanging Lived Experience as Peer Knowledge

Everyday experiences formed a shared, practical reference point. Descriptions ranged from *“I’m still lifting and running, and that’s feels great”* (PA, 2023) or *“I tried following exercise videos on YouTube, but it was too much for my body to handle,*”(PA, 2023) to feeling ‘*glued to a chair’*. (PA, 2023). Food experiences were shared just as openly, including *“This morning I ate a bowl of chips and salsa and queso at 8:00 in the morning!”* (Cravings, 2023). Humour was frequently deployed to cope with societal pressures and physical limitations, such as joking about a ‘*pregnancy Olympics*’ where ‘*putting on shoes’* (PA, 2022) was a major competitive event.

#### 1.3 Reciprocal Encouragement and Gratitude

The forum operates as a cyclical, community-building environment where users who receive reassurance frequently express deep gratitude and extend reciprocal emotional support.

Functioning as a support mechanism, these peer exchanges generate mutual motivation and reshape attitudes toward gestational body image. Community interactions prompted behavioural and cognitive shifts, with users stating, *“you are giving me some serious motivation to take my exercise routine serious and not be a sofa potato all day”* (PA, 2023), and “*I’ve realised with the help of you lovely folk that baby needs me healthy not skinny*” (PA, 2023). Furthermore, encountering shared experiences offered reassurance and mitigated feelings of isolation, leading to expressions of validation such as, *“it’s been so nice reading your stories and definitely brings me reassurance to know I’m not the only one being a couch potato”* (PA, 2021).

### Theme 2: Decision-Making Under Uncertainty

Those who post on Reddit navigated systemic information gaps, conflicting provider advice, and aggressive supplement marketing.

#### 2.1 Seeking Information Beyond Healthcare

Delays in accessing early antenatal care prompted users to seek interim advice online:

"it seems mad that you don’t get a GP appointment as soon as you find out you’re pregnant" **– (**PA, 2023). While waiting for appointments, Reddit became an interim source of practical guidance:*“My obgyn and ultrasound appointment are next week. Just want any advice until I get to speak to her”* - Supplements, 2021. Users compared experiences around food, weight, symptoms, and other pregnancy concerns, asking questions such as *“when did most ppl see the majority of weight gain-1st trimester, 2nd, or 3rd”(*GWG, 2023) and whether others had *“thrown caution to the wind with all the food no nos”* (Food safety, 2024).

Some users also expressed frustration with perceived gaps in maternal health knowledge: *“it doesn’t feel like they really have enough science on anything female let alone childbirth for us to have true guidance!”(*Prenatal anxiety, 2022). This uncertainty encouraged efforts to distinguish evidence from myths, including questions such as *“I heard it’s not safe to eat any processed meats… Is this true…?”* (Food safety, 2022) and requests for *“some kind of guide or app or resource that explains to me what’s safe week by week”*(PA, 2022).

Peer information seeking extended across diet, supplements, mental health, and physical activity. Users asked, *“How are you dealing with gaining too much weight?”(*GWG, 2023), *“do you have any recommendations on which prenatals to take*?”(Supplements, 2022)*“How do you guys calm yourself down from thinking the worst?* (Prenatal anxiety, 2024), and *“Is there such thing as safe exercises during pregnancy?”* (PA, 2021). More experienced exercisers also sought advice tailored to their routines: *“I’m an avid runner… Any other running Mama’s out there?”*(PA, 2020). Discussions further explored how lifestyle behaviours might affect pregnancy and birth outcomes. Users asked whether fitness provides a *“better womb in which to grow in”* (PA, 2019) whether exercise influences delivery, and how diet or weight relates to infant health: *“what your baby’s birth weight was and how healthy your pregnancy diet was”* (GWG, 2022) and *“Has obesity caused issues with your pregnancy?”* (GWG, 2021).

#### 2.2 Whose Expertise Counts?

Users weighed provider directives against bodily signals and peer experiences. Provider advice was challenged when perceived as outdated: *“I asked my dr but she gave me outdated advice about keeping my HR under 140”* (PA, 2020). Skepticism also extended to standard pregnancy recommendations, particularly when exercise or weight targets seemed unrealistic. One user questioned recommendations for 10,000 daily steps in late pregnancy, while another challenged advice that she should gain only 15 pounds because it *“seems kind of impossible based on all of the changes that occur during pregnancy”* (GWG, 2024). Users also compared professional guidance with the experiences of family, friends, and other mothers, sometimes finding peer perspectives more reassuring: *“Experienced moms at work:”This is your first and you’ve only gained 45lbs? You’re doing good! I gained 100lbs with my first*” (GWG, 2022). For some, previous negative healthcare experiences contributed to more explicit distrust:*“I have been misdiagnosed and given incorrect information so many times over the years by doctors, so I don’t take what they say as the end-all, be-all”* (GWG, 2022).

Weight and nutrition advice were particularly contested when users felt recommendations did not reflect their individual circumstances. Pregnant users questioned provider assessments of weight gain and expressed frustration with generic dietary advice delivered without discussion of their existing habits: *“Today, I was told to cut back carbs, to not have any soda or sugar, eat more protein - no discussion was had to ask what my food consumption looks like to begin with”* (GWG, 2022). Some posters recognized that providers *“come from a place of love and concern”* (GWG, 2024). When external recommendations conflicted with lived physical experience, bodily cues often became an alternative source of authority. Users described responding to hunger, comfort, and physical capacity rather than strictly following prescribed limits: *“I feel like I should listen to my body and just eat whatever I want”* (Hunger, 2022) and *“I feel fine running and listen to my body when I need to slow down*” (PA, 2022).

#### 2.3 Conflicting and Contradictory Guidance

The data illustrate that contradictions between healthcare providers, clinical guidelines, and social media algorithms generate confusion regarding sleep positions, exercise safety, and caloric restrictions. For example, one user noted, *“My obstetrician has approved me to keep exercising at the gym”* (PA, 2018), whereas another wrote, *“An OB told me any exercise besides walking was “bad for baby”* (PA, 2023). This friction was also evident in sleep guidance: *“An NP told me as long as I’m still comfortable it’s fine, but the OB said now that I’m 20 weeks I should be sleeping on my left side only”* (Sleep, 2021). Such systemic variance in clinical settings is compounded by conflicting online information, burdening users with the responsibility of auditing a chaotic mix of lifestyle rules, internet trends, and unresolved scientific debates. Users expressed extreme exhaustion over the *‘non-stop yes and no’* encountered across search engines and forums. Ultimately, users reported scouring websites only to *‘proceed to confuse myself’,* thinking that public guidelines surrounding exercise, caffeine consumption, and gestational weight benchmarks remain polarized.

#### 2.4 Balancing Guidance with Personal Needs and Perceived Risk

The findings reveal that pregnant individuals do not passively accept obstetric prohibitions; instead, users engaged in situational risk appraisal that balances official guidelines against emotional well-being, somatic needs, and localized logic. Indulging in cravings was frequently associated with feelings of intense autonomy and emotional comfort, with users reporting, *“This is the most craving-led day I’ve had to date and I have no regrets,”* (Cravings, 2021) or declaring: “*You’re supposed to STEAM your roast beef before eating??? No. I refuse…”* (Food safety, 2018).

### Theme 3: The Emotional Burden of Maternal Responsibility

Concerns regarding fetal safety contributed to heightened vigilance and feelings of guilt.

#### 3.1 Fear and Anxiety Around Pregnancy

Users routinely express maternal guilt when physiological symptoms prevent adherence to idealized lifestyle guidelines. Everyday choices are weaponized by self-doubt, as users fear that imperfect habits will harm the fetus: *“I feel awful!!! I’m scared my baby isn’t getting enough nutrients! … I can’t help but feel like I’m failing at taking care of my body”* (Hunger, 2022) and *“Sleeping on my back is a no no because of the anxiety that’s been planted in my head…”*(Sleep, 2019). Furthermore, GWG is rarely a neutral clinical metric. While some users express anxieties about feeling unready for parenthood, weight stagnation triggers panic over fetal starvation: *“I’m 15 weeks pregnant and my weight hasn’t budged… I’m a FTM* (first-time mom) *and really worried that I’m not gaining enough”* (GWG, 2023).

#### 3.2 Guilt, Self-Blame, and Maternal Responsibility

Maternal guilt acts as a psychological burden, with users comparing their biological realities to idealized pregnancy standards. Limitations beyond their control, and occasional departures from guidelines were internalized as personal failures that endangered the fetus: *“I messed up this baby’s development already…*” (Supplements, 2023) and “*I’m a terrible mom”* (Food safety, 2023). Additionally, medical labeling caused notable distress. Discovering epidemiological weight classifications on charts was experienced as a personal judgment by some users, overshadowing positive clinical indicators: *“I had my anatomy scan on Wednesday and everything went great, baby’s great, but I saw on my chart, the sono guy wrote that I was obese and it killed me to see that”* (GWG, 2022).

#### 3.3 Body Image and Weight-Related Identity

Some users reported that clinicians aware of patients’ disordered eating histories issued restrictive dietary mandates or ignored requests for blind weigh-ins, triggering distress: *“I mentioned to the person taking the appointment that I didn’t want to know my weight, she said no problem. So I stepped on the scales and she immediately told me, then told me my BMI. She said she wasn’t concerned but inside I felt sick”* (GWG, 2021). For users recovering from disordered eating, the rapid bodily changes of pregnancy coupled with medical surveillance threatened their psychological progress. The necessary discontinuation of psychiatric medications intensified this vulnerability: *“I never had a diagnosed ED, but I went through years of really detrimental restriction followed by exercise as punishment… I have spent so long working to undo those old mentalities… but every time I go to the doctor I am just feeling looked down on for my weight”* (GWG, 2022) and *“I struggle with binge eating and before getting pregnant had maintained a healthy weight for over 2 years. I have been off my meds since 12 weeks and cannot restart until baby is born”* (GWG, 2023).

### Theme 4: Embodied Knowledge in Practice

The theme captures how pregnant individuals actively adapt, negotiate, and survive the physical volatility of gestation by developing intuitive lifestyle strategies that balance clinical recommendations with their daily physiological and psychological realities.

#### 4.1 The Body’s Unpredictability

The users’ posts suggest a high level of physical volatility across gestation, where nausea, extreme appetite swings, food aversions, and pelvic pain forced them into ‘survival mode’. Users reported a lack of transition between symptoms: *“…does anyone else either feel sick to their stomach or starving like there’s no in between, I’m only a few weeks in”*(Hunger, 2020). When nausea and aversions became severe, eating was dictated entirely by keeping food down rather than meeting clinical targets: *“It was purely survival mode and I hated all food. I lost weight! I was throwing up multiple times daily for about 17 weeks”* (Hunger, 2023). Furthermore, Reddit users described disrupted sleep and mobility: *“I’m so tired of not getting rest and waking up 6 times a night because if I move it hurts”* (Sleep, 2020).

#### 4.2 The Shifting Palate

The findings highlight a transformation in sensory preferences during gestation, characterized by intense cravings that prompt immediate and sometimes costly behavioural responses: *“Oh, I’m ‘order 1 ice cream from cold stone through door dash at 9pm and pay 16 dollars because I can’t be bothered to drive to get it’ weeks along”* (Cravings, 2021). To manage these overwhelming desires, individuals described cycling between periods of high indulgence and corrective dietary phases, with one user noting, *“But I just want you to know I have an actual food hangover today! It’s gonna be salads, porridge and green smoothies from here on out….until the next craving binge!”*(Cravings, 2021). To better navigate these physical shifts, some users utilize the digital space to crowdsource practical nutritional strategies: *“Well what I have found is this symptom has improved greatly by reducing my meal size and eating more often…”* (Hunger, 2021).

#### 4.4: Adapting Recommendations to Lived Reality

Rather than strictly adhering to rigid protocols, pregnant individuals dynamically adapt clinical guidelines to fit their physiological limitations, with some users prioritizing fitness routines for emotional regulation: *“But I work out to stay healthy but most importantly for my mental health”*(PA, 2021). To minimize risks, users modify routines based on intuitive cues and utilize support devices for sleep “*I have a U-shaped pregnancy pillow which I use to stop me actually falling onto my back…”*. (Sleep, 2020). This active adaptation extends to dietary restrictions, where users adopt harm-reduction strategies over caffeine consumption: *“I always thought I’d fully quit but today I went out and bought decaf”* (Coffee, 2022). While some develop practical risk-management protocols to safely consume banned foods such as *“heating the meat in microwave first, and then also broiling my sandwich”* (Food safety 2023), the overwhelming volume of restrictions or forbidden foods leads others to abandon the rules in favour of intuitive desires: *“There are so many things pregos are not allowed to eat I don’t even care any more. If I want a sandwich I eat it. If I want coffee everyday I drink it.”* (Cravings, 2020).

### Theme 5: Social Pressure and Autonomy

Users actively resisted external surveillance and weight-focused clinical judgment.

#### 5.1 External Surveillance &

Pregnant individuals navigate a dense matrix of social observation, where their actions and dietary choices are constantly subjected to policing by strangers: “*A girl whose never been pregnant (why are they the ones?!) Starts guilting me for having coffee…”* (Caffeine, 2019). This behavioural regulation also extends into intimate relationships, where partners or other family members act as primary gatekeepers of safety rules based on digital literature or generational expectations. Users report experiencing this surveillance almost immediately:*“I’m so barely pregnant (4 weeks) and I told my husband yesterday. He’s really excited… but he’s already started laying down The Rules™ about what I can and can’t eat/drink”*(Caffeine, 2021). In addition, unsolicited comparisons add to this friction: *“Especially when my MIL says when I was pregnant I never gained any weight like, ok”* (GWG, 2023). Furthermore, this surveillance is heavily shaped by cultural backgrounds. Users reported struggling to balance medical guidelines against ingrained hospitality and dietary norms: *“But coming from the southern states, soda is a breakfast lunch and dinner thing”* (Caffeine, 2021).

#### 5.2 Provider Judgment

Formal clinical encounters were experienced as sites of external surveillance. Users detailed distressing appointments where clinical staff weaponized weight metrics and made assumptions, prompting some to report a loss of trust in their providers: *“I am so sorry for this, but I am starting to absolutely hate doctors”* (Caffein, 2022). Users reported feeling judged for their weight, noting:*”The entire appointment centered on my weight and he kept repeating my weight and shaking his head”* (GWG, 2023). This surveillance contributes to a broader perception of weight-centred prenatal care, where users think that clinicians treat BMI as the primary risk indicator: *“I feel like doctors here just pinpoint everything to being overweight if your BMI is high”* (GWG, 2021). Instead of accepting the criticism, some users actively research guidelines to contest practices: *“I just done some research (NHS) and it looks like the doctor was in the wrong… So no I am not morbidly obese I’m just pregnant”* (GWG, 2021). Ultimately, these challenging interactions can weaken the therapeutic alliance, prompting individuals to seek alternative providers to protect their well-being: *“I’m not planning on going back to her, and rescheduled with my regular doctor”*(GWG, 2021).

#### 5.3 Resistance, Autonomy, and Reclaiming Agency

The data illustrate that pregnant individuals actively develop strategies to navigate surveillance and maintain their personal autonomy. Users expressed frustration regarding unsolicited public advice, using the forum to reassert their decision-making capabilities: *“First of all, buddy, don’t tell me shit based off your outdated ‘information’ you’ve heard passed on through generations, and second of all, I have a doctorate, … You think I haven’t looked into whether I can have coffee?”* (Caffeine, 2022) and use sarcasm to critique the stereotypes: *“Like, yeah, pregnant women lose all common sense and ability to control themselves in the face of cravings. I was totally planning on downing a 6-pack and lighting up a Marlboro, Karen”* (Cravings, 2023). This pushback extends to challenging imposed physical limitations, with users firmly stating, *“I’m pregnant, not disabled”* (PA, 2022). In clinical settings, this self-advocacy translates into a demand for healthcare transparency, as users question administrative protocols that complicate their choices: *“its actually shocking that women lack so much autonomy and choice that me thinking about not being weighed is such an issue for so many people. Your autonomy and my autonomy is important”* (GWG, 2023).

## Discussion

This study combined BERTopic modeling of 15 years of Reddit posts with reflexive thematic analysis to examine how pregnant individuals navigate health behaviours and gestational outcomes online. Previous research has shown that women seek pregnancy information online. By using a long timeframe and a mixed-methods design, this study goes further and maps the broader informational ecosystem. Posts about immediate gestational milestones such as fetal movement (Rank 1; 5,454 posts), account for the largest share of forum volume overall. However, health outcomes remain a major recurring focus of digital peer interaction, led by gestational weight gain (Rank 14; 2,241 posts).

Taken together, the topic modeling and thematic analysis suggest that forum activity reflects unmet clinical needs. The most active topics are not randomly distributed. The heavy concentration of posts about GWG, prenatal supplements, cravings, and prenatal exercise points to where women may experience the heaviest emotional burden and scrutiny. The prominence of weight gain fits with earlier work showing that gestational weight gain is a source of considerable uncertainty, anxiety, and social scrutiny for pregnant women, and that it is among the topics they look up online most often (Sasidharan et al., 2025; Willcox et al., 2015; Zhou et al., 2023). Caffeine intake, sleep position, and food safety each generated hundreds of posts, which further reflects the fragmented and often conflicting guidance women encounter, prompting them to seek reassurance and normalization from peers rather than, or in addition to, their clinicians (Djouma Nembot et al., 2026).

Consequently, the most discussed topic areas are likely the ones where women most need to make sense of things together and lean on what their bodies tell them. The five qualitative themes that follow show how users navigate these high-priority topics.

Our first theme showed that Reddit functioned primarily as a space for collective sense-making. The reassurance, validation, and normalization users sought mirror what online pregnancy communities are known for. Emotional and appraisal support are consistently the most frequently reported benefits, and they help ease isolation, low mood, and anxiety. Women turned to peers specifically to confirm that unexpected experiences like hunger, missed supplements, and weight gain were normal, echoing evidence that normalization is a central driver of forum use (Lee & Lee, 2022; Morris et al., 2026). Sharing everyday details as "peer knowledge" reflects women’s preference for learning from other women’s first-hand experience, and their use of forums as a source of clinical information between appointments (Ellis & Roberts, 2020). The encouragement and gratitude users offered one another also illustrate the mutual, respect- and shared-responsibility-based nature of peer support, where giving and receiving helps fosters self-esteem and a sense of belonging (Morris et al., 2026).

Across its five sub-themes, Theme 2 captures how women make pregnancy decisions when the answers are not clear. Turning to informal and online sources aligns with evidence that women seek information beyond formal care when it feels delayed, inaccessible, rushed, or insufficiently woman-centred, filling perceived gaps in unbiased maternal health information (Ellis & Roberts, 2020; Morris et al., 2026). The question of "whose expertise counts" reflects the tension between professional guidance and lived, embodied authority. Women accept clinical advice selectively, weigh it against peer and bodily cues, and disengage when it feels vague, outdated, or generic, consistent with literature showing that trust and individualized, respectful communication determine whether provider guidance is followed (Vedam et al., 2019).

Conflicting and contradictory guidance adds to the difficulty. It is a well-documented quality problem in digital maternal health information, where women report overwhelming volume, inaccuracies, lack of scientific evidence, and pervasive contradictions. Algorithms also passively expose users to unvetted material that can both reassureand heighten fear (e.g., pregnancy-loss content) (Ellis & Roberts, 2020; Li et al., 2025). Even a simple, well-established recommendation, such as taking a daily prenatal supplement, becomes less clear once women must interpret it against a crowded market of competing formulations and promotional claims (Caro & Fast, 2020). Finally, balancing clinical guidance against personal comfort and symptoms reflects situational risk appraisal rather than simple non-adherence. This active cost-benefit analysis highlights the need for counseling that respects maternal autonomy while still prioritizing the most critical, evidence-based precautions (Caro & Fast, 2020; Rockliffe et al., 2021).

Theme 3 captures the emotional burden of maternal responsibility that pervades these discussions. Fear and anxiety surrounding fetal safety, food risk, sleep position, and weight gain contribute to pregnancy-related anxiety. This includes worry about infant health, body appearance, and one’s adequacy as a parent (Escañuela Sánchez et al., 2022). That anxietyin turn fuels guilt and self-blame built around the "good mother" ideal, whereby socially constructed norms of motherhood and unrealistic expectations generate distress when lived reality falls short, and health behaviour change in pregnancy becomes structured around adopting a "good mother" role (Harrison et al., 2020). Finally, concerns about body image and weight-related identity are consistent with evidence that appearance-related sociocultural pressure drives body dissatisfaction, and that insensitive, weight-focused care can reactivate vulnerability to disordered eating (Sollid et al., 2021).

Theme 4 shows that embodied knowledge, more than information alone, shapes health behaviour in practice. Users described an unpredictable body, with nausea, extreme hunger, food aversions, and disrupted sleep, and they adapted recommendations to cope. This finding aligns with syntheses showing that lived physical experience, not knowledge alone, drives dietary, activity, and sleep behaviour in pregnancy. As pregnancy progressed, users described pain and increasing fatigue, which contributed to declining physical activity across gestation (Newson et al., 2022). Barriers to dietary supplement adherence are nausea, taste and smell aversion, and forgetting to take supplements. These potentially modifiable barriers may be addressed through alternative formulations and reminder strategies, supporting guideline recommendations for daily prenatal vitamin supplementation. More broadly, across supplementation,physical activity, sleep, caffeine, diet, and food safety, pregnant individuals actively regulated their own behaviour rather than simply complying or not complying with recommendations. Behaviour change in pregnancy is therefore best understood as an ongoing negotiation between clinical guidance and lived physical reality. This scenario reinforces calls for flexible, individualized counselling rather than prescriptive, rule-based messaging (Forbes et al., 2018).

In Theme 5, women described being watched, by the people around them and by their providers, over what they eat, drink, and do, and how much they weigh. This feeling of surveillance reflects the influence of subjective norms and social approval on pregnancy health behaviour, with family and cultural context acting as both support and stressor. Provider judgment and weight-centred pushback align with evidence that women report the lowest decision-making autonomy when they feel pressured, dismissed, or rushed, and that weight stigma erodes trust and prompts them to switch providers. In response, women reasserted control over their bodies and care decisions, reclaiming a sense of agency and self-determination. This aligns with most women’s strong preference to be lead decision-makers in their care and underscores the importance of autonomy as a core element of respectful, person-centred maternity care, particularly when guidance feels coercive or poorly justified (Souza et al., 2020; Vedam et al., 2019).

## Implications for Maternal Healthcare

- **1. Earlier Access to Evidence-Based Information:** Antenatal care models should provide accessible, evidence-based digital resources immediately upon pregnancy confirmation to bridge the gap before the initial clinical appointment.
- **2. Empathetic, Non-Stigmatizing Communication:** Clinicians must move away from rigid, weight-centric surveillance and BMI-shaming. Clinical conversations should focus holistically on modifiable health behaviors rather than scale weight alone.
- **3. Flexible, Person-Centred Guidance:** Guidelines for physical activity, diet, and supplementation should be presented as flexible targets that adapt to physical symptoms (nausea, fatigue, joint pain). Provider counselling must explicitly respect patient autonomy and embodied decision-making.

## Strengths and Limitations

A key strength of this study is its extensive 15-year longitudinal scope, which provided a broad dataset capturing the evolving health concerns of a large digital pregnancy community. Additionally, the pseudo-anonymous nature of Reddit creates an "incognito" environment that may reduce social desirability bias compared to traditional in-person interviews or clinical settings allowing unfiltered discourse regarding sensitive topics such as gestational weight gain, maternal guilt, and deviations from dietary guidance.

Nevertheless, several limitations should be considered when interpreting these findings. First, the use of de-identified social media data precluded the collection of demographic and clinical metadata (e.g., age, parity, socioeconomic status, and clinical history), limiting the transferability of the results. Relatedly, platform selection bias is inherent to Reddit, whose user base skews toward younger, English-speaking, and digitally literate demographics, potentially underrepresenting non-Western or digitally marginalized populations. Methodologically, analyzing static archival text prevented iterative qualitative probing to clarify users’ rationale or track longitudinal outcomes. Finally, because these data reflect self-reported narratives and subjective interpretations rather than objectively verified medical records, the findings lack clinical corroboration.

## Conclusions

By pairing computational BERTopic modeling of 15 years of online discourse with reflexive thematic analysis, this study highlights a meaningful gap between standardized clinical guidance and the lived physical realities of pregnant individuals. Pregnant women rely on digital peer communities to normalize physical struggles, evaluate conflicting advice, and exercise bodily autonomy. Bridging this gap requires maternal healthcare systems to deliver information and care that is evidence-based, flexible, empathetic, and individualized.

## Data Availability

All data produced in the present work are contained in the manuscript

